# Is there a difference in the satisfaction levels between professionals who have participated and those who have not participated in the TeleUTIP telemedicine project regarding their engagement in tele-education activities?

**DOI:** 10.1101/2024.01.16.24301368

**Authors:** Hilda M. R. Moleda Constant, Vanessa Cristina Jacovas, Maria Eulália Vinadé Chagas, Maria Cristina Cotta Matte, Emanuele König Klever, Luciane Cunha, João Ronaldo Mafalda Krauzer, Taís de Campos Moreira, Felipe Cezar Cabral

## Abstract

**Objective:** The aim of this study is to introduce a distance education program and assess the satisfaction of health professionals.

**Methods:** A cross-sectional study was conducted in five public hospitals across different regions of Brazil. A total of 171 health professionals participated in a tele-education activity involving discussions of complex cases in a Case-Based Learning (CBL) format. Employee satisfaction was measured using the translated questionnaire Tele-Education Satisfaction Assessment Instrument (IAS-TE).

**Results:** The mean satisfaction score for participants was 4.6 on a 5-point Likert-type scale. Participants engaged in the telemedicine project (35.5%) reported significantly higher satisfaction (p<0.001) compared to their non-participating peers (n = 149, 64.5%). The IAS-TE exhibited a Cronbach’s alpha of 0.73, indicating good reliability, and demonstrated good homogeneity (KMO = 0.76, Bartlett’s test of sphericity = p<0.00).

**Conclusion:** Participants in the proposed activity expressed high levels of satisfaction. The findings suggest that satisfaction levels are elevated among professionals involved in the telemedicine project. The proposed satisfaction assessment instrument (IAS-TE) exhibits robust psychometric qualities and is suitable for evaluating other distance education activities. Future perspectives involve gaining a deeper understanding of strategies to enhance satisfaction for professionals who do not engage in activities using advanced technology.

## INTRODUCTION

The use of health technologies has increased significantly after the coronavirus pandemic. These tools, previously seen as a complement to improve health care, have become an emerging need in the field. The applicability of these resources is currently not limited to assistance and reach different areas, such as the training of health professionals. Different technologies can be used as tools in this context. With regard to assistance, telemedicine is an alternative that has been successful in supporting situations of social isolation and lack of specialized professionals^1–3^. As for professional training, tele-education has been widely used in the health area, mainly for continuing medical education^4^. Some studies have proven that education technologies are highly effective in supporting the learning process^5,6^.

There are countless tele-education models available to assist in this process, which differ largely in terms of the technology and media employed^4^. Nowadays, the use of tele-education has become a fundamental tool for professionals to maintain learning even at a distance. One increasingly popular model is case-based learning (CBL), which, in medical education, is based on the discussion of clinical cases. This methodology aims to prepare students for real-world care by linking theory to practice through the application of knowledge to clinical practice^7^. In the intensive care unit (ICU) setting, CBL has been associated with both improvements in clinical outcomes and cost savings^8^. Furthermore, this learning model is suitable for helping students become active learners, increasing knowledge and understanding, creating empathy, and promoting decision making^9,10^. The use of CBL in training healthcare providers also seeks to prompt changes in practice.

Usually, to evaluate the impact of using these technologies, studies of continuing education practices have reported the perceived satisfaction of participants with the activities performed^11,12^. Satisfaction is one of the most important outcomes measured in tele-education^13^. Indeed, it has been cited as one of the five pillars of quality in distance education^14^. Many studies have assessed satisfaction in activities that use the CBL model^15,16^. However, evaluation of the use of this model in tele-education and associated with telemedicine is a gap in the literature. Seems to be interesting to understand what aspects can influence the satisfaction of participants in these activities, thus it will be possible to plan better this practice. Besides, due to the expansion of the use of technologies, there is an immediate need for studies on the application of these tools. Therefore, the objective of this study was to present a distance education program and evaluate the satisfaction of health professionals. For evaluating the satisfaction of the health professionals, we take into account who participated and those who did not in a telemedicine project and in a distance education activity. Also, there are no brief instruments in Brazil that evaluate satisfaction in this format of activity. Even though it is an essential aspect, satisfaction is sometimes evaluated through questionnaires with unknown psychometric properties, limiting the validity of the results^17,18^. Therefore, secondly, an analysis was realized of the psychometric properties of a brief satisfaction assessment instrument (Tele-Education Satisfaction Assessment Instrument-*Instrumento de Avaliação da Satisfação em Tele-Educação-IAS-TE*) that will be available for other studies on tele-education.

## METHODS

### Delimitation

A cross-sectional study was carried out to assess satisfaction and to analyze the psychometric properties of the IAS-TE.

### Settings and participants

The study took place as part of the project “Qualification of Assistance in Pediatric Intensive Care by Telemedicine - TeleUTIP”, which aims to offer, via telemedicine, continuing care education and specialized medical care for PICUs, in places where distance and the lack of specialized professionals is a difficulty. The project’s proponent team is in a control room, where they carry out daily tele-rounds with the teams of the participating hospitals (remote centers) that are in the PICUs of their hospitals and have a telemedicine cart capable of connecting the teams to assist and outline better clinical practices. Four health institutions participated in the study, in addition to the proposing team.

In addition to the tele-rounds, the project aims to contribute to the continuing education of the professionals involved in the rounds and the other professionals in the units, this occurs through discussions of complex cases in the format of CBL-based. To achieve this goal, cases previously attended by these professionals were discussed in order to provide an opportunity for reflection on what would be the best care plan for these patients. In order to contribute to the discussions, specialist physicians in the topic were invited to participate in the discussion case. Additionally, it is understood that this program can serve as feedback from the proposing center to the teams participating in the project. The invitation to participate in the CBL-based was made for all professionals of the PICUs that are part of the TeleUTIP project and not exclusively for the professionals who were directly involved with the tele-rounds. In general, the discussions of complex cases were composed of the same groups of participants but they were invited again in each meeting. All participants were invited to answer the satisfaction questionnaire in each meeting. Therefore, the survey could be answered more than once by the same participant in different meetings on different topics. The participants are therefore divided into two groups:

<u>TR + CBL:</u> composed for health professionals who participated in the tele-round and education activities of the discussions of complex cases.

<u>CBL:</u> composed for health professionals who participated only in education activities of the discussions of complex cases.

### TeleUTI distance education program

The infrastructure required was: a portable speaker and medium-range microphone connected to a computer, a multimedia projector, and an appropriate room or other venue for video conferencing. The cases that would be discussed were selected continuously during the sessions. The professional, responsible for the presentation, selected the case that in his professional practice was a challenge. We opted to use a script for conduct activity and a model of the abstract to case and presentation because several studies describe that it is essential for the professionals to adhere to this activity that the cases discussed are as close as possible to the reality of the professional’s practice^10,12,13^. It is important to note that patient privacy was guaranteed by anonymizing private patient data.

To ensure standardization of presentation materials, all participating facilities were sent a roadmap for case preparation (Supplementary Material 1) and a presentation template. In addition to this material, participants were requested to choose two published scholarly articles to support their case discussions.

Twelve synchronous CBL-based sessions were held, lasting 40 minutes each. With each session, the participating hospital responsible for presenting the case rotated. The sessions were held between July 2019 and March 2020. Within each session, the case presentations were all aided by audiovisual resources and lasted 15 minutes. After each presentation, a discussion of the case was mediated by specialists in the topic, who had been previously invited to participate. Furthermore, participants were given additional educational materials (journal articles) to support the learning process. The discussion groups consisted of physicians, pediatric residents, and a multidisciplinary team. After each activity, the IAS-TE was administered to the participants. Those who were affiliated with the project’s host hospital were asked to complete it on the spot, while participants from the other centers received the questionnaire via e-mail at the end of the activity.

### Tele-Education Satisfaction Assessment Instrument-IAS-TE

The instrument used for the activity was adapted from the existing literature^19^. It is a brief (4-item), easy-to-answer questionnaire. The items were designed so as to cover four factors: content, technical aspects, duration, and interaction with participants. The decision was made to have only four items in the adapted version due to the need for quick administration and to ensure participant adherence at this stage of the activity. The full version of the instrument is given in Supplementary Material 2. The items address the following topics:

1. content: quality of the topic, relevance to clinical practice, level of understanding of the topic;
2. technical/operational issues: perception of audio, video, and connection quality;
3. length of the activity;
4. interaction with participants (how the speaker addressed questions during the session).

Answers to these questions were scored on a 5-point Likert scale, where 0 = not at all satisfied and 5 = totally satisfied. The sample comprised 171 health professionals.

### Data analysis

As satisfaction is one of the fundamental variants for evaluating distance education activities, an assessment of the IAS-TE structure was carried out in order to ensure of the data quality in this study and in order to make the use of the instrument feasible in future studies. The structure of the IAS-TE was reviewed in terms of internal consistency through exploratory factor analysis (EFA) (Supplementary material 3).

All other data collected in this study were subjected to univariate statistical analyses. Quantitative variables are described as means and standard deviations or medians and interquartile ranges as appropriate. Categorical variables are expressed as absolute and relative frequencies. Bivariate analyses were conducted through Student’s *t*-test for independent samples. For purposes of analysis, participants were divided into two groups— complex case discussion + tele-rounding and complex case discussion alone— to test for differences in satisfaction between the participants involved in actual patient care rounds versus those involved in the Grand Rounds-type activity only.

All analyses were carried out in the Statistical Package for the Social Sciences (SPSS), version 18.5. P-values <0.05 were considered statistically significant.

### Ethics approval

This study was approved by the Research Ethics Committee of Moinhos de Vento Hospital. All participants signed an Informed Consent Form.

## RESULTS

### Participant satisfaction

A total of 231 professionals from 5 different hospitals across Brazil participated in the meetings of case discussions. Of these, 171 (73.5%) completed the satisfaction surveys, 109 participants were from the proposing institution and 62 were from the other hospitals. Of the total sample of participants, 103 were physicians, 118 were doctor residents, and 10 were nurses and/or other members of the multidisciplinary team. 69 participants (30%) were male. Among the respondents, 149 (64.5%) were from the CBL group and 21 (35.5%) were from the TR + CBL group.

All institutions participating in the project received an invitation to case discussion and were allowed to forward this invitation to local health professionals. Therefore, not all the professionals were involved in the tele-rounds. Figure 1 summarizes the study protocol. The mean satisfaction score was 4.6 (out of a maximum 5 points). The outcome of each case discussion is shown in Table 1. When comparing satisfaction with participation in case discussions between professionals who were involved in tele-rounds and those who were not, we found that the mean overall satisfaction of professionals involved in tele-rounds was 4.85, versus 4.58 among those who were not involved in this activity (p<0.01) (Table 2). The mean satisfaction per group in each IAS-TE question was shown in figure 2.

**Table 1.** Result of each complex case discussion (CBL).

| Topic | Number of participants | Number of satisfaction survey respondents | Overall satisfaction* |
| --- | --- | --- | --- |
| Bilateral diaphragmatic paralysis | 17 | 17 | 4.5 |
| Omphalocele | 25 | 18 | 4.3 |
| Acute renal failure requiring dialysis | 24 | 20 | 4.6 |
| Septic shock and diabetic ketoacidosis | 23 | 16 | 4.7 |
| Necrotizing pneumonia | 10 | 10 | 4.5 |
| Spina bifida | 20 | 17 | 4.5 |
| Toxic epidermal necrolysis (SJS/TEN) with chronic complications | 14 | 13 | 4.7 |
| Sexual differentiation disorder (bilateral cryptorchidism) | 25 | 13 | 4.4 |
| An infant with jaundice, liver failure, and bicytopenia | 16 | 8 | 5 |
| Anomalous origin of left coronary artery: Takeuchi repair | 27 | 19 | 4.7 |
| An infant with a fungal infection | 12 | 8 | 4.9 |
| An infant in close contact with a tuberculosis patient since birth | 18 | 11 | 4.8 |

**Table 2.** Mean satisfaction for each IAS-TE question.

| | | Satisfaction Mean ± dp | p* value |
| --- | --- | --- | --- |
| Dimensions | TR + CBL Group<br>n = 21 | CBL Group<br>n = 150 |  |
| Content | 4,90 ± 0,30 | 4,70 ± 0,56 | 0,07 |
| Operational Technical Issues | 4,76 ± 0,43 | 4,22 ± 0,84 | 0,00* |
| Time of Duration | 4,90 ± 0,30 | 4,73 ± 0,51 | 0,02* |
| Interaction Participants | 4,86 ± 0,39 | 4,60 ± 0,62 | 0,05* |

\*p ≤ 0.005

**Figure 1.**
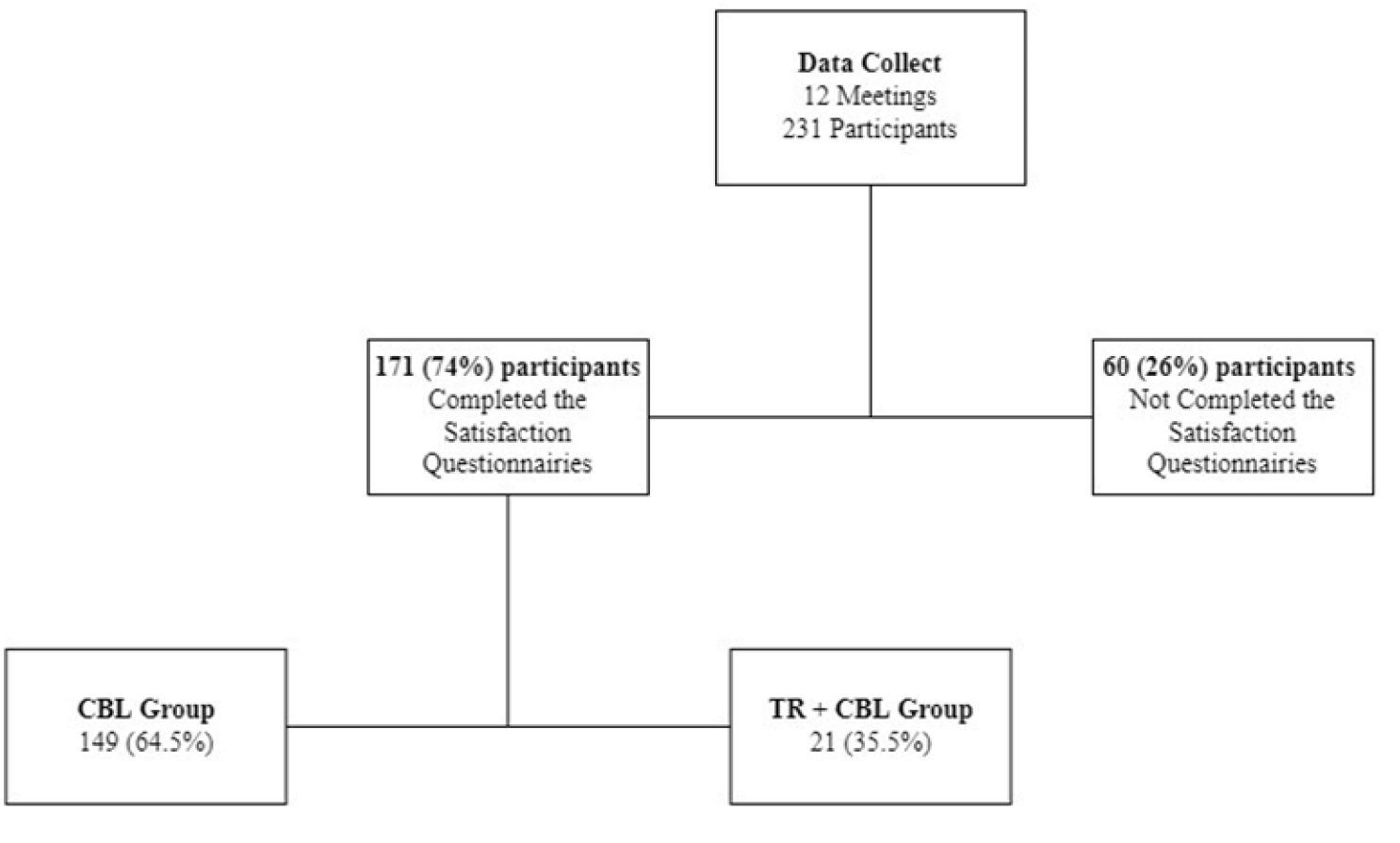
Study protocol.

**Figure 2.**
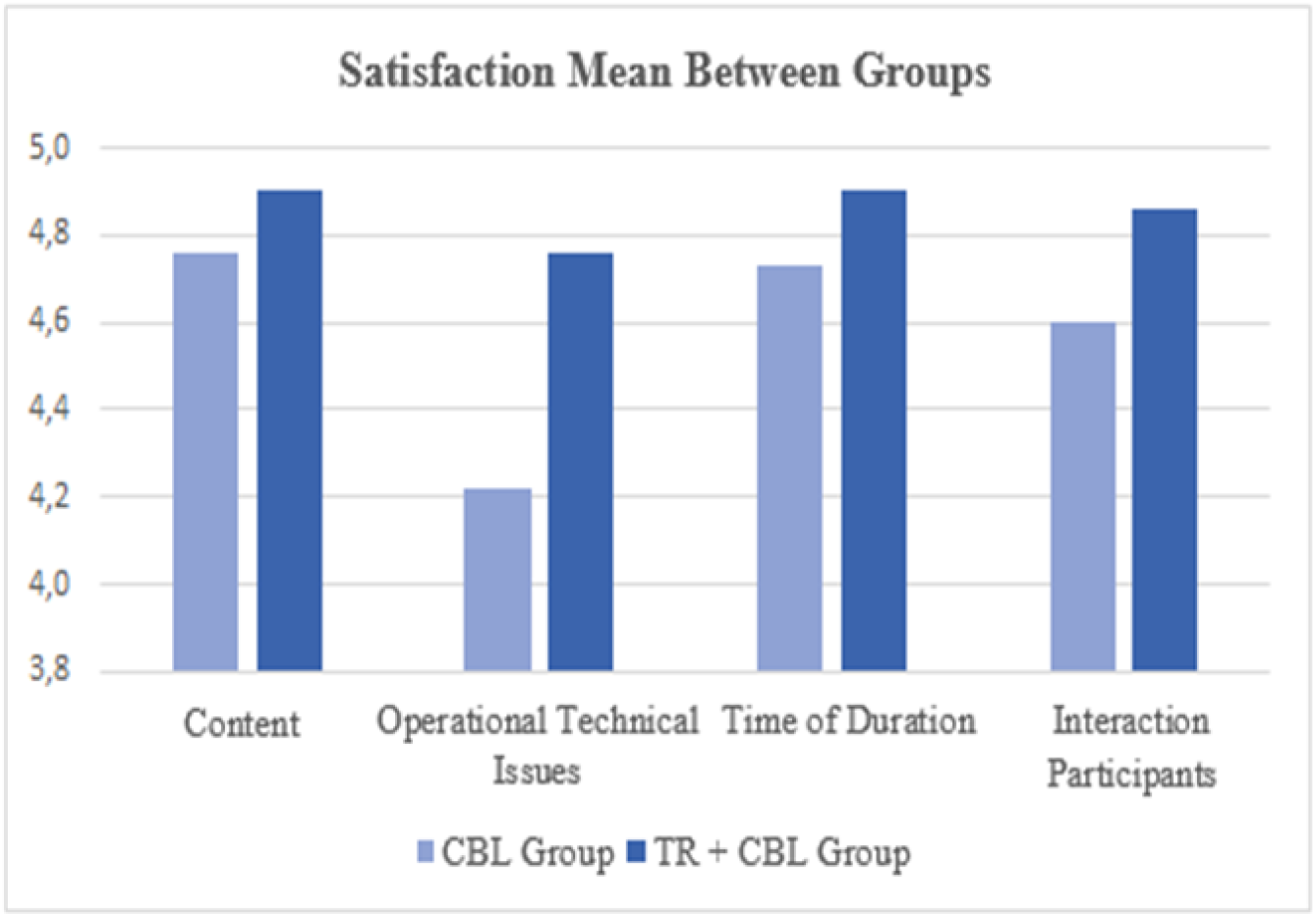
Mean satisfaction per group in each IAS-TE question.

### Psychometric properties of the IAS-TE

Cronbach’s alpha was 0.73, which is considered acceptable, and the total variance explained was 57%. EFA yielded a moderate KMO index (0.76), and Bartlett’s test of sphericity showed p<0.001 and a chi-square value of 145.38 (df = 6). Confirmatory factor analysis resulted in the extraction of a single factor (Supplementary material 4).

## DISCUSSION

The work presented a distance education program joined with tele-round activities that can be considered a learning option that results in good satisfaction of the participants. The average satisfaction of PICU staff members who participated in complex case discussions was high. Several studies have reported high satisfaction with educational activities based on this method^10^. However, few studies have assessed whether there are differences in satisfaction with learning through the CBL model among professionals who participate in actual patient care activities, such as clinical rounds, compared to those who do not. Knowledge of this aspect can help plan optimal CME activities for professionals who are involved in direct patient care. When we analyzed satisfaction separately by professionals involved versus not involved in pediatric tele-rounds, there was greater satisfaction among professionals who participated in both activities (Grand Rounds-style complex case discussion and patient care rounds). Combining different activities such as patient care with Grand Rounds discussions may therefore lead to better participant satisfaction and superior learning, and reflect more directly on an improvement in quality of clinical care. In the hospital setting, education using real patient cases helps students formulate a more integrated and multidisciplinary learning experience. It helps involve everyone in problem-solving, builds critical thinking skills for the analysis of complex issues, and increases satisfaction and confidence in knowledge acquisition^13^.

The CBL model has been shown to produce highly satisfied and engaged professionals, perhaps due to the more in-depth learning made possible by this model^11,13^. Additionally, it is well known that satisfaction can lead to greater motivation and engagement in learning activities^10^. This is the main advantage of knowing which learning models promote greater satisfaction and healthcare education quality in participants.

Additionally, it was possible to verify the validity of the satisfaction instrument used, which makes it reliable for use in similar activities. It is worth highlighting that the psychometric properties of assessment instruments should always be evaluated in the population to which they will be administered, thus ensuring that the instrument will really gauge what it is designed to measure. In respect, to possible limitation for be brief instrument, although the literature reports that instruments composed of few items tend to have a low alpha^20^ value, the same did not occur with our questionnaire. Despite its brevity, the IAS-TE presented acceptable internal consistency, that is, reliability^21^. In addition, the results obtained demonstrate that the EFA can prove a good correlation between the items and a percentage of explained variance consistent with those described in the literature^20,22^. In other words, the evidence suggests that this instrument measures what it was designed to measure. It fulfills its main purpose, which is to determine the satisfaction of the participants with the key aspects of the activity (content, technical issues, length, facilitator interaction with the participant) while being easily and quickly administered.

The limitations of this study include I) The scale used was a 5-point Likert-type scale, so the statistical differences can be considered discrete in the items; II) Participants from TR + CBL group is much lower from the CBL group. This was because it was an open invitation activity for professionals; III) Absence of sociodemographic data of the sample, due to the lack of time for all answers, the most relevant ones were chosen; IV) The participants’ learning was not measured. However, other instruments as brief as the IAS-TE have been shown to evaluate the satisfaction construct adequately and yield excellent results^19,21^. With these limitations, our results are not possible to generalize to other educational activities at a distance.

## CONCLUSIONS

The work presented a distance education program that can be considered a learning model that deliver satisfaction for participants. In this study, professionals who participated in patient care activities (tele-rounds) as well as in a Grand Rounds-type activity of complex case discussion reported higher satisfaction levels than those who participated in case discussions alone.

The satisfaction assessment instrument designed for this activity, the IAS-TE, was found to have good psychometric qualities. It is especially suitable for use in situations where rapid questionnaire administration is essential and to assess satisfaction with dynamic distance education activities, such as tele-education.

This work is the first step for evidence of the new practices in distance education associate the telemedicine. Interesting avenues for future research include evaluation of knowledge acquired and of the impact of this activity on clinical practice, as well as measurement of whether greater clinical confidence is associated with greater satisfaction. These perspectives demonstrate the wide potential for further exploration of this topic

## Data Availability

All data produced in the present work are contained in the manuscript.

## ACKNOWLEDGMENTS

The authors thank the Research Support Center-NAP of the Hospital Moinhos de Vento for the technical-scientific support and Guilherme Carey Fröhlich, biomedicine student.

## FUNDING

The author(s) disclosed receipt of the following financial support for the research, authorship, and/or publication of this article:

The data presented was obtained in partnership with the Ministry of Health of Brazil, through the Institutional Development Support Program of the Unified Health System (PROADI-SUS).

