## Supplemental Data 1 for "Is there a difference in the satisfaction levels between professionals who have participated and those who have not participated in the TeleUTIP telemedicine project regarding their engagement in tele-education activities?"

### Supplemental data 1: Roadmap for case vignette.

#### ROADMAP FOR CASE VIGNETTE:

#### COMPLEX CASE DISCUSSION

These *Complex Case Grand Rounds* will take place between Hospital Moinhos de Vento (HMV) and the remote hospitals participating in the TELEUTIP project. The aim of this Continuing Medical Education activity is to support the training of Pediatric Intensive Care Unit (PICU) staff at the participating hospitals (PICU staff, Pediatrics residents, and Pediatric Critical Care residents) through presentation and discussion of complex cases with the HMV team. In addition, it aims to provide for the cross-institutional exchange of information on diagnostic and management approaches, as well as expose residents and multidisciplinary PICU team members to challenges and diversity in care settings and systems they might not otherwise encounter. The Rounds will follow a structured model (similar that employed in the partnership between HMV and Johns Hopkins International), which is described in further detail below.

The timetable of activities will be set by prearrangement between the participating hospitals. At each meeting, one hospital will be in charge of presenting a case proposal. The participating professionals will be the PICU coordinators of the participating institutions, the coordinators and staff physicians of the telemedicine centers, and the corresponding multidisciplinary teams. To assist in organization, each hospital will have a medical officer designated as its focal point for the Rounds. Specialists will be invited to speak at the Rounds according to the needs of the selected case. At HMV, participants will be informed of the location in advance; at the telemedicine centers, participants will be sent a link 24 hours in advance.

Structure of case presentation (must be prepared by a physician/resident + focal point):

**1**- Selection of a complex case by the hospital whose turn it is according to the rotation.

**2**- Preparation of a brief PowerPoint presentation following the provided template. This presentation will be shared to all during the stream.

**3**- Writing a brief report (example below), to the mailed to the list of participants up to 96 hours before the date of the presentation. One or two published articles relevant to the topic (to support the discussion) should be attached with the report.

"As part of the HMV TELEUTIP Project, we have scheduled the next complex case discussion for [MONTH DAY, YEAR], at TIME, presented by [NAME OF HOSPITAL]. The case will be presented by [NAME OF RESIDENT]. The case concerns a patient with [DIAGNOSIS] and

[COMORBITIES/COMPLICATIONS]. This discussion will provide participants with an opportunity for [XXXX] (e.g.: evidence-based debate), and may involve [SPECIALISTS] (e.g.: cardiologists, pulmonologists, and a multidisciplinary team). Finally, the focus of the discussion should be [FOCUS OF DISCUSSION] (e.g.: "the management of acute decompensations, including plans for continuity of care – optimization of ventilation strategies, duration of tracheostomy, catheter use, medications…)"

**4**- It is suggested that, in addition to the participants, [SPECIALISTS, MULTIDISCIPLINARY TEAM MEMBERS (NURSES, NUTRITIONISTS, PHYSICAL THERAPISTS, PSYCHOLOGISTS)] be invited as appropriate according to the features of the case. If the invited professionals are unable to participate, they are advised to appoint a colleague who can attend in their stead.

**5**- The presentation will be conducted by the pediatrics resident or pediatric critical care resident.

**6**- The recommended length of the presentation is 15 minutes (max. 5 slides) followed by a 45-minute discussion.

**7**- At the end of the case, a satisfaction survey and attendance sheet will be sent to the participants. Both must be completed and delivered to the focal point.

**Note:** All complex case presentations will be recorded. The footage shall remain on file with the HMV coordinators and can be used in future training activities for other teams participating in the project.
