## Supplemental Data 2 for "Is there a difference in the satisfaction levels between professionals who have participated and those who have not participated in the TeleUTIP telemedicine project regarding their engagement in tele-education activities?"

TELEUTIP PROADI SUS MOINHOS DE VENTO

In order to obtain information about your satisfaction with this activity, we would like you to answer the following questions on a scale of 0 to 5, where 0 = not at all satisfied and 5 = totally satisfied:

1. How satisfied are you with the content of the presentation (quality, relevance to clinical practice, and level of understanding)? *

*Mark only one oval.*

| | 0 | 1 | 2 | 3 | 4 | 5 | |
|---|---|---|---|---|---|---|---|
| Not at all satisfied | ◯ | ◯ | ◯ | ◯ | ◯ | ◯ | Totally satisfied |

2. How satisfied are you with your experience from a technical standpoint (audio, video, and connection)? *

*Mark only one oval.*

| | 0 | 1 | 2 | 3 | 4 | 5 | |
|---|---|---|---|---|---|---|---|
| Not at all satisfied | ◯ | ◯ | ◯ | ◯ | ◯ | ◯ | Totally satisfied |

3. How satisfied are you with the length of the presentation? *

*Mark only one oval.*

| | 0 | 1 | 2 | 3 | 4 | 5 | |
|---|---|---|---|---|---|---|---|
| Not at all satisfied | ◯ | ◯ | ◯ | ◯ | ◯ | ◯ | Totally satisfied |

4. Regarding the interaction with the participants (doubts clarified during the session): *

*Mark only one oval.*

| | 0 | 1 | 2 | 3 | 4 | 5 | |
|---|---|---|---|---|---|---|---|
| Not at all satisfied | ◯ | ◯ | ◯ | ◯ | ◯ | ◯ | Totally satisfied |
