## Supplemental Data 3 for "Is there a difference in the satisfaction levels between professionals who have participated and those who have not participated in the TeleUTIP telemedicine project regarding their engagement in tele-education activities?"

**Data analysis of IAS-TE**

In order to carry out the evaluation of the structure of the IAS-TE was reviewed in terms of internal consistency through exploratory factor analysis (EFA). Factor analysis was justified by the assumptions of large sample size, multivariate normality, linearity, and correlation between variables[1,2]. The statistical assumption of multivariate normality was verified by plotting a histogram of the four metric variables that make up the questionnaire, using the values obtained from skewness and kurtosis tests as well as the Kolmogorov–Smirnov test. To verify the assumption of linearity, scatter plots of pairs of variables were analyzed. To test for correlation between variables, an analysis of reliability was also conducted to assess internal consistency by Cronbach's alpha and homogeneity, followed by exploratory factor analysis (EFA) with varimax rotation. Eigenvalues greater than 1 were accepted[3,4]. The Kaiser–Meyer–Olkin (KMO) test measures were interpreted as per Hutcheson and Sofroni[5] and Bartlett's test of sphericity[6].

**Supplemental data references**


1. Knapp RR and Comrey AL. Further Construct Validation of a Measure of Self-Actualization. Educational and Psychological Measurement 1973;33(2):419–425; doi: 10.1177/001316447303300225.

2. Tabachnick BG, Fidell LS and Ullman JB. Using Multivariate Statistics. Seventh edition. Pearson: NY, NY; 2019.

3. Harman HH. Modern Factor Analysis. University of Chicago Press; 1976.

4. Kaiser HF. The Varimax Criterion for Analytic Rotation in Factor Analysis. Psychometrika 1958;23(3):187–200; doi: 10.1007/BF02289233.

5. Anonymous. The Multivariate Social Scientist. 2022. Available from: https://uk.sagepub.com/en-gb/eur/the-multivariate-social-scientist/book205684 [Last accessed: 8/3/2022].

6. Kaiser HF and Rice J. Little Jiffy, Mark Iv. Educational and Psychological Measurement 1974;34(1):111–117; doi: 10.1177/001316447403400115.
